# A 6-Item Diagnostic Screener for Childbirth-Related PTSD

**DOI:** 10.64898/2026.03.05.26347629

**Authors:** Alon Bartal, Tal Elhasid Felsenstein, Hadas Allouche-Kam, Elli C. Dassopoulos, Mary C. Lee, Andrea G. Edlow, Scott P. Orr, Sharon Dekel

## Abstract

Posttraumatic stress disorder (PTSD) after traumatic childbirth is a serious maternal morbidity that affects about 20% of women following medically complicated deliveries. Yet it often goes undetected in perinatal care, in part because brief screening tools suitable for busy obstetric settings are lacking. We developed and evaluated a brief screener derived from the 20-item PTSD Checklist for DSM-5 (PCL-5) to detect childbirth-related PTSD. Women with traumatic childbirth experiences completed the PCL-5 and the gold-standard Clinician-Administered PTSD Scale for DSM-5 (CAPS-5); depression symptoms were assessed with the Edinburgh Postnatal Depression Scale (EPDS). Bootstrap resampling with LASSO regression identified the PCL-5 items most strongly associated with PTSD, and Firth logistic regression was used to estimate diagnostic accuracy. A statistically derived 6-item version of the PCL-5 (PCL-5-R6) showed excellent discrimination against clinician diagnosis (AUC = 0.95; 95% CI, 0.89–1.00). A cut-off score of 7 yielded high sensitivity (0.96) and good specificity (0.83), with overall diagnostic efficiency of 0.86. The PCL-5-R6 correlated only moderately with EPDS scores (ρ = 0.53), indicating that depression screening alone does not reliably detect PTSD. This brief tool can offer a valid and practical approach for routine postpartum PTSD screening.

## Introduction

Reports of traumatic childbirth often involve a perceived threat to the life or physical integrity of the mother and/or her infant, or the possibility of serious injury. These reports are frequently associated with medically complicated deliveries, including moderate to severe obstetrical complications or unexpected medical interventions (e.g., unscheduled Cesarean delivery)^1–4^. In the U.S., an estimated 30% of births entail various degrees of complication, with a minority (1.8%) resulting in severe maternal morbidity^5, 6^.

Posttraumatic stress disorder (PTSD) stemming from a traumatic childbirth experience, also referred to as childbirth-related PTSD (CB-PTSD), is an under-recognized postpartum mental health condition^1, 7^. CB-PTSD has a prevalence of 5.9 % for all deliveries and is estimated between 18.5% to 41.2% for women who have had a medically complicated/traumatic delivery^2, 3, 8–10^.

CB-PTSD symptoms are evoked by reminders of the traumatic childbirth and manifest in constant re-experiencing of the traumatic childbirth, avoidance of trauma reminders, alterations in cognitions and mood, and hyper-reactivity^1, 3, 7^. The infant can serve as a reminder of the traumatic childbirth and evoke maternal distress^1,7,11^. Unresolved CB-PTSD symptoms can result in impairment in maternal bonding and breastfeeding problems that can have negative effects on the child during a critical period of early child development^3, 7, 12–15^.

A first step in treatment and prevention of postpartum mental conditions is timely, efficient, and low-burden screening that can be readily implemented in routine perinatal care assessments during the postpartum period. Because postpartum mental health symptoms may be attributed to postpartum depression, and because of limited provider awareness of CB-PTSD, women at risk for CB-PTSD may be underdiagnosed, misdiagnosed, and under-treated^1, 16, 17^.

The American College of Obstetrics and Gynecologists (ACOG) recommends screening for mental health conditions using validated measures^18^. The gold standard for PTSD diagnosis is the Clinician-Administered PTSD Scale (CAPS-5), which entails a structured interview for assessing the presence and severity of PTSD symptoms and requires up to an hour of a trained professional’s time^19^. Patient self-report instruments can be used as a screen for PTSD before an in-depth evaluation is undertaken. One of the most widely used screening scales is the PTSD Checklist for DSM-5 (PCL-5)^20^. It shows strong test-retest reliability and excellent agreement with clinician-diagnosed PTSD^20,21^. The diagnostic usefulness of the PCL-5 Checklist for the postpartum population has been documented for PTSD following traumatic childbirth^8^. The Checklist assesses the 20 DSM-5 PTSD symptoms and takes about 10 minutes to complete, which may be considered too long or burdensome for the purpose of early perinatal care. Development of a very brief screening tool that minimizes patient burden while efficiently identifying probable CB-PTSD cases would be of value.

The objective of the reported study was to develop and evaluate an optimally shortened version of the PCL-5 in a sample of women who had experienced a traumatic childbirth, which included women with a clinically confirmed diagnosis of CB-PTSD. The study measured the contribution of each of the 20 PCL-5 items against the clinician CB-PTSD diagnostic status derived from the CAPS-5 and selected 6 items that best predicted the clinician diagnosis. The diagnostic accuracy of the shortened PCL-5 scale was compared to that of the full scale.

## Materials and Methods

### Study Population

The study included 107 women who had a traumatic childbirth experience. All had obstetrical complications and/or underwent an unscheduled medical intervention(s) during labor or delivery. Table 1 presents detailed demographic and obstetric information of the study cohort. [**Table 1**. insert here]

**Table 1.** Demographics and obstetrical information of the study sample.

| Variable | *n (%) / M (SD)* |
| --- | --- |
| Maternal age at delivery | 32.62 (4.30) |
| Parity |  |
| Primiparas | 73 (68.22%) |
| Multiparas | 34 (31.78%) |
| Obstetrical Complications/Conditions |  |
| Hemorrhaging/excessive blood loss | 27 (25.23%) |
| Hysterectomy | 4 (3.74%) |
| Preeclampsia/Eclampsia | 17 (15.89%) |
| Blood transfusion/infusion | 10 (9.35%) |
| Gestational diabetes | 10 (9.35%) |
| Infection | 7 (6.54%) |
| Fetal intolerance | 44 (41.12%) |
| Failed labor progression | 37 (34.58%) |
| Arrest of descent | 6 (5.61%) |
| Placenta abruption | 4 (3.74%) |
| Placenta previa/Low-lying placenta | 8 (7.48%) |
| Cord prolapse | 3 (2.80%) |
| Nuchal cord | 4 (3.74%) |
| ICU admission, maternal | 2 (1.87%) |
| Mode of delivery |  |
| Vaginal | 25 (23.36%) |
| Vaginal assisted | 15 (14.02%) |
| Scheduled Cesarean | 4 (3.74%) |
| Unscheduled or emergency Cesarean | 63 (58.88%) |
| Gestation age (wk) | 39.22 (1.84) |
| Neonatal medical complications | 15 (14.02%) |
| Demographic information at psychometric testing |  |
| Maternal age (yrs) | 33.24 (4.24) |
| Years after delivery (median) | 0.2 |
| Marital status |  |
| Married or living with partner | 102 (95.33%) |
| Single | 5 (4.67%) |
| Education |  |
| Formal college degree or higher | 97 (90.65%) |
| No formal degree | 10 (9.35%) |
| Race |  |
| White | 81 (75.70%) |
| Asian/Asian-American | 11 (10.28%) |
| Black/African-American | 3 (2.80%) |
| Other | 12 (11.22%) |
| Ethnicity |  |
| Hispanic/Latinx | 13 (12.15%) |
| Not Hispanic/Latinx | 94 (87.85%) |

Note: Medical Information is derived from electronic medical records and/or patient self-reporting; demographic information pertains to patient's status at study assessment. Neonatal complications: Medical complications requiring neonatal intensive care unit (NICU) admission. ICU: Intensive care unit admission. Two patients were missing data for maternal age at delivery and study assessment, and percentages were calculated based on valid values. Years after delivery range between 0.08 and 9.76. n = 107.

All patients met the formal Diagnostic Statistical Manual fifth edition (DSM-5) A criterion for PTSD that addresses threat or potential threat to life or serious injury, experienced or perceived, as it pertained to them and/or their newborn. Patients completed the PCL-5 and were also evaluated during a structured clinical interview using the CAPS-5 to assess for CB-PTSD presence and symptom severity. All patients provided written informed consent before the study assessments. The reported sample is derived from a larger study of the mental health sequela of childbirth; all studies were approved by the Mass General Brigham (MGB) Research Committee and performed in accordance with its relevant guidelines and regulations.

Recruitment methods primarily included hospital announcements and targeted enrollment using information from obstetric medical records indicative of medically complicated childbirth.

### Measures

Self-reported PTSD symptoms, related to the patient’s childbirth experience, were measured with the PCL-5 Checklist^20^. It lists the 20 symptoms of PTSD based on the DSM-5 that are rated by the patient on Likert scales (Not at all (0); A little bit (1); Moderately (2); Quite a bit (3); and Extremely (4). The PCL-5 has demonstrated strong psychometric performance with high reliability and validity observed in various trauma-exposed samples^20, 21^. The Checklist previously has been used in postpartum women following traumatic childbirth; the total symptom severity score on the Checklist was strongly associated with the clinician assessment^8^.

The presence and severity of PTSD symptoms related to traumatic childbirth were assessed by a clinician using the CAPS-5^19^. The CAPS-5 includes 30 items that correspond with the DSM-5 PTSD criteria and takes 45 to 60 minutes to administer. A PTSD diagnosis requires the presence of exposure to a traumatic event (DSM-5 criterion A) and endorsement of PTSD symptoms in the past month encompassed within 4 symptom clusters: intrusions, avoidance, alterations in cognitions and mood, and hyperarousal^22^.

Self-report depression symptoms were assessed using the Edinburgh Postnatal Depression Scale (EPDS)^23^. It is the recommended tool to screen for perinatal depression during routine perinatal care in medical settings in the U.S^18, 24^.

#### Analysis

We identified key items from the 20 PCL-5 items with the strongest predictive contribution to the CAPS-5 CB-PTSD diagnosis status using a three-step process (Appendix 1). Our approach employed bootstrap resampling for model training and evaluation, with each iteration generating a new training set while the remaining observations served as the out-of-bag (OOB) testing set.

**Step 1:** We used LASSO regression to identify PCL-5 items most predictive of CAPS-5 CB-PTSD diagnosis.

**Step 2:** Using each bootstrap training set, we trained two adaptive models and a baseline model. **Step 3:** Model performances were evaluated on the OOB testing set to assess whether a reduced set of PCL-5 items could achieve diagnostic performances comparable to the full 20-item PCL-5. Mean sensitivity, specificity, and AUC were calculated with 95% confidence intervals (CIs). OOB predictions were pooled across iterations to generate Receiver Operating Characteristic (ROC) curves, and differences in AUC were tested using DeLong’s test^25^ for correlated ROC curves. The best-performing model was defined as the one with a significantly higher AUC.

### Developed Models. Adaptive Model 1 – Reduced-Multi

A Firth-penalized logistic regression in which the PCL-5 items selected in Step #1 were entered as individual predictors of CAPS-5 CB-PTSD diagnosis. This multivariable approach preserves item-level detail but may be unstable in small samples and is less practical as a brief screening tool. **Adaptive Model 2 – Reduced-Sum:** To address instability, the PCL-5 items selected in Step #1 were aggregated into a single summed score predictor of CAPS-5 CB-PTSD diagnosis. A summed (total) score decreases model complexity and provides a stable, interpretable index for evaluating diagnostic performance. **Baseline Model 3 – Full-Sum:** Uses the total score of all 20 PCL-5 items to predict CAPS-5 CB-PTSD diagnosis. Note. Adaptive Models #1 and #2 use LASSO-selected PCL-5 items within each resample, and therefore the item set can vary across bootstrap iterations.

To derive a fixed set of reduced PCL-5 items, we plotted *Stability* (item selection frequency in Step #1 across bootstrap iterations) against *Contribution* (median drop in OOB AUC when permuting each item while holding others constant). Items were classified into four groups: (1) strong and stable, (2) strong, (3) moderate, and (4) weak (Appendix 1). Only items in Groups #1 and #2 were retained to construct the final fixed reduced model. Optimal cutoffs for the fixed reduced and full 20-item PCL-5 models were identified using Youden’s J statistic^26^, and classification performances were compared with McNemar’s test^27^. We report performance metrics including Overall Diagnostic Efficiency (ODE).

Finally, we assessed the association between the fixed reduced PCL-5 total score and Edinburgh Postnatal Depression Scale (EPDS) total score using Spearman’s rank correlation (ρ), which does not assume normality and is suitable for summed questionnaire scores.

## Results

Among the 107 participants 27 met CB-PTSD diagnostic criteria, and 80 did not. The mean PCL-5 sum score based on 20-items was 18.58 (range 0-57). Two participants each had 1 missing PCL-5 value, and one participant had 2 missing PCL-5 values. Two participants each had 1 missing EPDS value. These were inferred via mean participant score on the non-missing values of the participant. Two participants without EPDS value were discarded for the EPDS analysis.

Table 2 presents the mean sensitivity, specificity, and AUC for the three models across 1,000 bootstrap resamples, with 95% confidence intervals: Adaptive Model 1 (Reduced-Multi), Adaptive Model 2 (Reduced-Sum), and the Baseline Full-Sum model.

**Table 2.** Bootstrap performance of PCL-5 models for predicting CAPS-5 CB-PTSD diagnosis.

| Model | AUC<br>(95% CI) | Sensitivity<br>(95% CI) | Specificity<br>(95% CI) |
| --- | --- | --- | --- |
| Model 1. Reduced-Multi | 0.862<br>(0.644–0.971) | 0.714<br>(0.333–1.000) | 0.822<br>(0.667–0.962) |
| Model 2. Reduced-Sum | 0.915<br>(0.831–0.980) | 0.803<br>(0.500–1.000) | 0.816<br>(0.667–0.962) |
| Model 3. Full-Sum | 0.923<br>(0.848–0.980) | 0.851<br>(0.600–1.000) | 0.785<br>(0.607–0.940) |

Note. Values represent mean performance across 1,000 bootstrap replications with corresponding 95% confidence intervals (CIs). Full-Sum = PCL-5 20 item total score; Reduced-Sum = PCL-5 selected items total score not limited to 6 items; Reduced-Multi = selected items set with multivariable model. AUC = Area Under the Curve.

The Reduced-Sum model outperformed the Reduced-Multi model (AUC = 0.92 vs 0.86; DeLong *p* = 0.03) and performed comparably to the Full-Sum model (AUC = 0.92; DeLong *p* = 0.23), indicating that the summed selected PCL-5 items offers better performance than item-level modeling while maintaining predictive performance comparable to the full 20-item scale (Table 2, Figure 1; Appendix 2). [**Table 2** and **Figure 1** insert here.]

**Figure 1.**
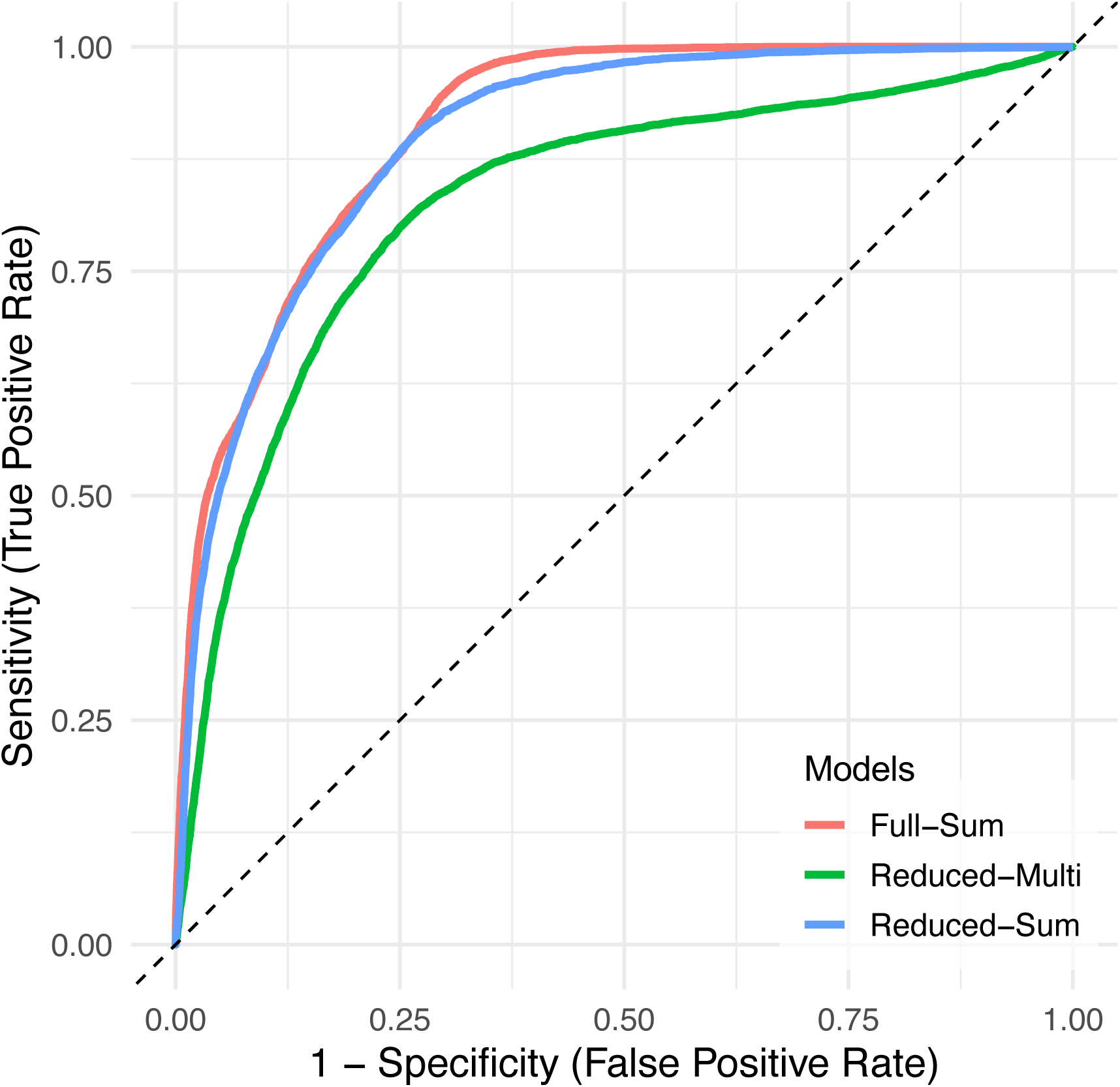
ROC curves of the pooled out-of-bag (OOB) predictions for the two Adaptive models and the full 20-item PCL-5 baseline model. The ROC curves of the Reduced-Sum and Full-Sum models intersect, and their overall discrimination did not differ significantly (DeLong test, *p* = 0.23), suggesting comparable average performance with no clear dominance across all thresholds. ROC curves are based on pooled OOB predictions across 1,000 bootstrap resamples. The Adaptive models used LASSO-selected PCL-5 items within each resample; consequently, the item set may vary across bootstrap iterations.

### Development of a fixed set of reduced PCL-5 items

Using the Stability and Contribution metrics, we classified the 20 PCL-5 items into four importance groups (Figure 2). (1) Strong and stable: *pcl_q10*, *pcl_q2*, *pcl_q7, pcl_q15*; (2) Strong: *pcl_q4*, *pcl_q9*; (3) Moderate: *pcl_q12*, *pcl_q17*, *pcl_q8*, *pcl_q14*, *pcl_q20*, *pcl_q19*, *pcl_q16*, *pcl_q13*, *pcl_q3, pcl_q18*; and (4) Weak: *pcl_q11*, *pcl_q5*, *pcl_q1*, *pcl_q6*. [**Figure 2**. insert here.]

**Figure 2.**
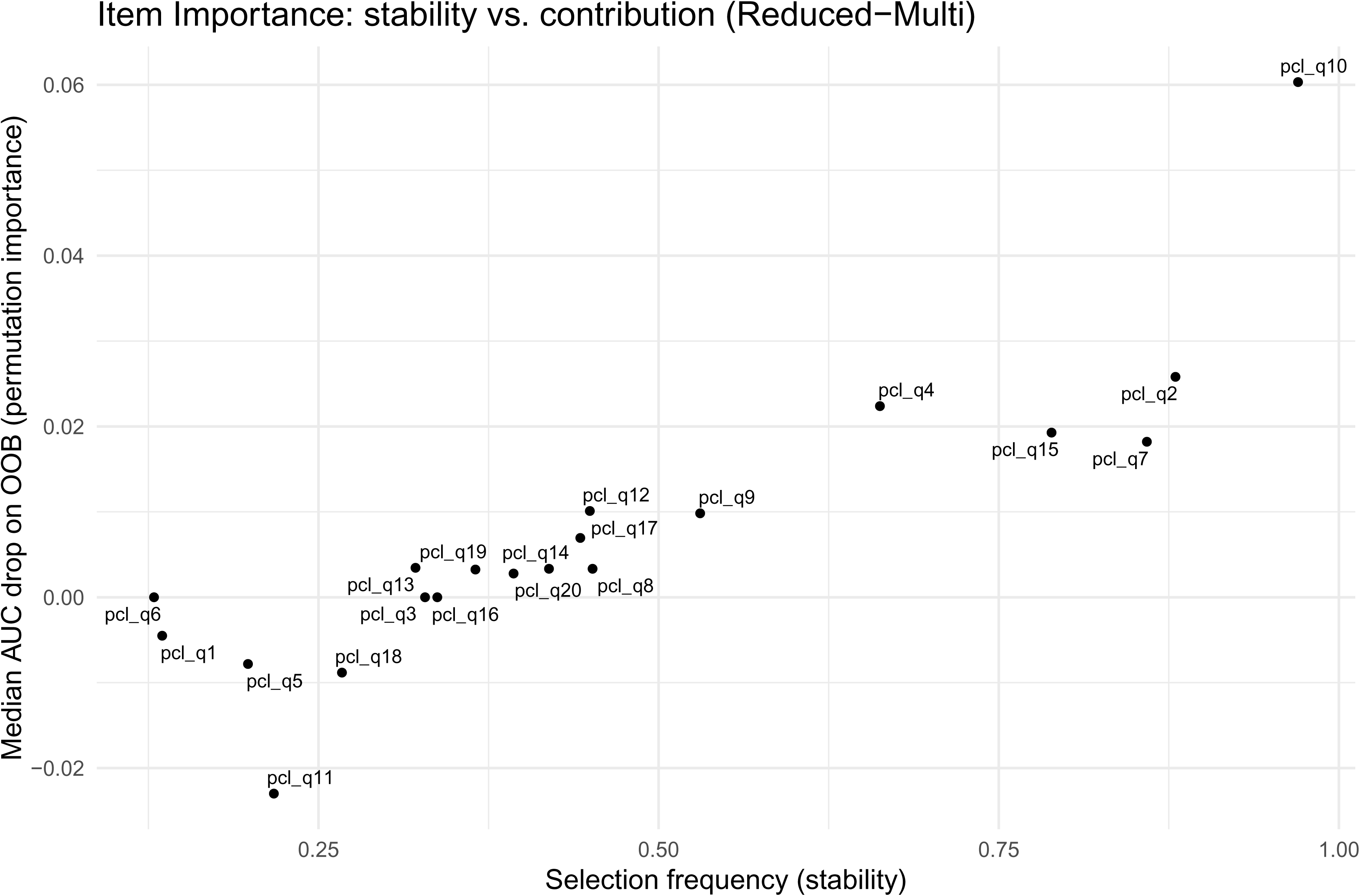
PCL-5 items importance across bootstrap resamples. The x-axis shows selection frequency (proportion of resamples in which an item was selected by LASSO). The y-axis shows the median out-of-bag (OOB) AUC drop when permuting the item while holding other selected items fixed. Items in the top-right quadrant are stable and high-contribution predictors. PCL-5 = PTSD Checklist for DSM-5.

We defined a fixed reduced model by summing the six items in Groups 1 and 2 {pcl_q10, pcl_q2, pcl_q7, pcl_q15, pcl_q4, pcl_q9}, denoted: PCL-5-R6 (Appendix 3). The PCL-5 R6 demonstrated strong diagnostic accuracy for CAPS-5 CB-PTSD status across cutoff values (AUC = 0.95, 95% CI: 0.89–1.00). A total score of 7 on the PCL-5 R6 maximized Youden’s J-statistic (0.78), providing the optimal balance between sensitivity (0.96), specificity (0.83), and overall diagnostic efficiency (ODE = 0.86) (Table 3). [**Table 3**. Insert here.]

**Table 3.** Diagnostic use of the Reduced PTSD Checklist for 6-items of the DSM-5 by cutoff value scores.

| Cutoff | Sensitivity | Specificity | PPV | NPV | J-statistic | ODE |
| --- | --- | --- | --- | --- | --- | --- |
| 6 | 0.963 | 0.788 | 0.605 | 0.984 | 0.750 | 0.832 |
| **7** | **0.963** | **0.825** | **0.650** | **0.985** | **0.788** | **0.860** |
| 8 | 0.852 | 0.863 | 0.676 | 0.945 | 0.714 | 0.860 |
| 9 | 0.852 | 0.913 | 0.767 | 0.948 | 0.764 | 0.897 |
| 10 | 0.778 | 0.938 | 0.808 | 0.926 | 0.715 | 0.897 |
| 11 | 0.630 | 0.963 | 0.850 | 0.885 | 0.592 | 0.879 |
| 12 | 0.481 | 0.988 | 0.929 | 0.849 | 0.469 | 0.860 |
| 13 | 0.407 | 0.988 | 0.917 | 0.832 | 0.395 | 0.841 |
| 14 | 0.370 | 0.988 | 0.909 | 0.823 | 0.358 | 0.832 |
| 15 | 0.259 | 0.988 | 0.875 | 0.798 | 0.247 | 0.804 |

Note. DSM-5: Diagnostic and Statistical Manual of Mental Disorders, Fifth Edition; NPV: negative predictive value; ODE: overall diagnostic efficiency; PCL-5: PTSD Checklist for DSM-5; PPV: positive predictive value; PTSD: posttraumatic stress disorder.
Cutoff values refer to PCL-5 (PTSD in regard to childbirth) summed (total) 6-item symptom severity score (PCL-5 R6). Cutoffs < 6 have a sensitivity of 1 and NPV of 1, whereas cutoffs > 15 have a specificity of 1 and a PPV value of 1.
Total PCL-5 R6 cutoff of 7 achieves optimal balances between sensitivity and specificity, identified by maximized Youden's J statistic.

Paired McNemar tests showed no significant difference between the PCL-5 R6 and the 20-item PCL-5 at their respective optimal cutoffs (χ² = 0.31, p = 0.58). For the full scale, a cutoff of 23 yielded sensitivity of 0.85, specificity of 0.84, and ODE = 0.83 (Table 4). [**Table 4**. Insert here.]

**Table 4.** Diagnostic use of the PTSD Checklist for 20-items of the DSM-5 by cutoff value scores.

| Cutoff | Sensitivity | Specificity | PPV | NPV | J-statistic | ODE |
| --- | --- | --- | --- | --- | --- | --- |
| 19 | 0.889 | 0.747 | 0.522 | 0.956 | 0.636 | 0.785 |
| 20 | 0.889 | 0.759 | 0.533 | 0.957 | 0.648 | 0.776 |
| 21 | 0.889 | 0.782 | 0.558 | 0.958 | 0.670 | 0.794 |
| 22 | 0.852 | 0.816 | 0.590 | 0.947 | 0.668 | 0.822 |
| **23** | **0.852** | **0.839** | **0.622** | **0.948** | **0.691** | **0.832** |
| 24 | 0.815 | 0.851 | 0.629 | 0.937 | 0.665 | 0.841 |
| 25 | 0.778 | 0.862 | 0.636 | 0.926 | 0.640 | 0.841 |
| 26 | 0.778 | 0.862 | 0.636 | 0.926 | 0.640 | 0.832 |
| 27 | 0.741 | 0.885 | 0.667 | 0.917 | 0.626 | 0.85 |
| 28 | 0.741 | 0.885 | 0.667 | 0.917 | 0.626 | 0.841 |
| 29 | 0.741 | 0.908 | 0.714 | 0.919 | 0.649 | 0.86 |
| 30 | 0.630 | 0.931 | 0.739 | 0.890 | 0.561 | 0.879 |
| 31 | 0.593 | 0.931 | 0.727 | 0.880 | 0.524 | 0.85 |
| 32 | 0.556 | 0.931 | 0.714 | 0.871 | 0.487 | 0.841 |
| 33 | 0.556 | 0.954 | 0.789 | 0.874 | 0.510 | 0.85 |
| 34 | 0.556 | 0.954 | 0.789 | 0.874 | 0.510 | 0.85 |
| 35 | 0.556 | 0.977 | 0.882 | 0.876 | 0.533 | 0.869 |
| 36 | 0.556 | 0.977 | 0.882 | 0.876 | 0.533 | 0.869 |
| 37 | 0.556 | 0.977 | 0.882 | 0.876 | 0.533 | 0.869 |
| 38 | 0.519 | 0.977 | 0.875 | 0.867 | 0.496 | 0.869 |

Note. DSM-5: Diagnostic and Statistical Manual of Mental Disorders, Fifth Edition; NPV: negative predictive value; ODE: overall diagnostic efficiency; PCL-5: PTSD Checklist for DSM-5; PPV: positive predictive value; PTSD: posttraumatic stress disorder.
Cutoff values refer to PCL-5 (PTSD in regard to childbirth) summed (total) 20-item symptom severity score. Cutoffs < 19 have a sensitivity of ~1 and NPV of ~1, whereas cutoffs > 38 have a specificity of 1 and a PPV value of 1.
Total 20-item PCL-5 cutoff of 23 achieves optimal balances between sensitivity and specificity, identified by maximized Youden's J statistic.

When analyses were limited to CAPS-5–positive cases (sensitivity) or CAPS-5–negative cases (specificity), differences between the PCL-5 R6 and the full 20-items remained non-significant (sensitivity: χ² = 1.33, p = 0.25; specificity: χ² = 0, p = 1.00). These findings indicate that the short six-item PCL-5 performs comparably to the full scale in both sensitivity and specificity.

Finally, the PCL-5 R6 total score showed a moderate positive correlation with the EPDS total score (Spearman’s ρ = 0.53, p < 0.001), supporting discriminant validity by indicating partial but not complete overlap between posttraumatic stress and depressive symptoms.

## Discussion

### Principal findings

This study identified a short, fixed set of six PCL-5 items (PCL-5 R6) that effectively screen for PTSD after traumatic childbirth. Using stability and contribution metrics, we found that these items, representing specific DSM-5 symptoms, can consistently captured the strongest signals for PTSD diagnosis. The resulting PCL-5 R6 showed excellent diagnostic accuracy when compared to a gold-standard clinical interview, while requiring far less patient and clinician burden. It performed comparably to the full 20-item scale. Importantly, its moderate correlation with the Edinburgh Postnatal Depression Scale (EPDS), highlights that depression measures alone cannot reliably detect childbirth-related PTSD. Together, these findings support the PCL-5 R6 as an evidence-based, efficient tool for identifying women at risk for PTSD in obstetric and postpartum settings.

### Results in the Context of What is Known

Existing PTSD screening tools for detecting PTSD related to traumatic childbirth have been largely developed in other trauma-exposed groups and often lack diagnostic validation in the postpartum population^63,81^. We examined the diagnostic utility of a short patient self-report assessment for detection CB-PTSD in high-risk women who had a medically complicated childbirth. No brief tool has been validated before. To this end, we developed an abbreviated instrument from the most commonly used screening tool for PTSD (PCL-5)^20^. We used statistical methods to identify key PCL-5 items that informed development of the short 6-item version. To determine diagnostic performance, we evaluated it in comparison to the traditional 20-item PCL-5 when benchmarked against the gold-standard, clinician-administered diagnostic assessment.

A few existing studies of individuals exposed to other forms of trauma support the diagnostic value of brief PCL-5 versions (of 4 and 8 items)^29–31^. Prior work has primarily used heuristic methods to inform shortened scales rather than statically derived approaches and has limited incorporation of the gold-standard clinician interview (CAPS-5) as the external diagnostic criterion. No brief versions of the PCL-5 have been validated for women who experienced birth trauma. Childbirth is an inherently unique experience, symptom profile, frequency, severity, salience and interpretation can vary depending on the type of trauma. As with other short PCL-5 versions, the scale developed in this study represents symptoms from each of the DSM-5 PTSD symptom clusters, supporting the formal conceptualization of a trauma-related disorder in the postpartum population.

### Clinical Implications

PTSD resulting from the experience of a traumatic childbirth remains underdiagnosed in routine perinatal care^7^. Currently, screenings in hospitals and clinics in the U.S. primarily target symptoms of depression using the EPDS^18^, which do not capture symptoms of PTSD. This leaves an unmet clinical need that requires attention. Women at high risk for CB-PTSD mostly represent those with medically complicated childbirth who are also at risk for experiencing serious short-and long-term physical complications^2, 3, 32^. ACOG warrants using validated tools for maternal mental health screening within the comprehensive 12 weeks postpartum visit using measures that are easily administered and scored^24^. Here, we provide evidence supporting an assessment that uses a reduced-item set of only 6 structured questions targeting specific DSM-5 PTSD symptoms.

The advantage of the short PCL-5 is that it allows for a brief assessment that is designed to save time but still retain strong diagnostic accuracy. Therefore, this version can be ideally suited for the busy outpatient clinic in which perinatal providers are required to complete a comprehensive obstetrics assessment and also screen for other mental health conditions^24^.

Availability of a brief screening tool with minimal patient and staff burden is often the determining factor of whether a condition will be assessed or not. The EPDS has offered universal screening for peripartum depression and has been widely adopted in clinics globally, resulting in a marked improvement in the detection of depression^18, 24, 33^. The availability of scalable and timely screeners for CB-PTSD will likely have similar clinical benefits. Because CB-PTSD, unlike peripartum depression, is not formally recognized in the DSM-5 (PTSD with a postpartum onset specifier is not listed), providers and patients may attribute symptoms to normal postpartum recovery or to signs of depression. Adopting a screening for CB-PTSD on a large scale can serve as the first step towards improving awareness of this maternal morbidity and reducing underdiagnosis and misdiagnosis.

### Research Implications

An important question that remains largely unanswered is whether CB-PTSD resembles the features of PTSD related to other forms of trauma. This understanding will likely require biological studies to reveal the disorder’s underlying mechanisms and inform novel screening methods rather than adopting measures that were originally developed in non-postpartum populations. The heavy reliance on patient self-report of their symptoms with no identified biomarkers to improve screening is common practice in psychiatry. Within these limitations, future studies are needed to replicate our findings and test the clinical usefulness of the 6-item version of the PCL-5 in larger postpartum samples of women who experience a traumatic childbirth and its acceptability in the clinic.

### Strengths and Limitations

We tested a short assessment for detection of PTSD in targeted patients with traumatic childbirth. The identified tool was developed using a statical approach and evaluated for diagnostic performance. We further used statistical techniques (resampling) to test the stability of the findings based on many random samples drawn from the data. Optimal clinical cutoffs were identified that could inform assessments for possible CB-PTSD. The developed scale was examined also in relation to the recommended measurement of postpartum depression symptoms performed in the clinic (EPDS) to highlight the unique contribution of an assessment specific to traumatic stress.

Several weaknesses of this study should be noted. They include the relatively small sample with confirmed CB-PTSD, the use of re-sampling within the existing dataset, and homogeneity of the study cohort. Prospective large-scale studies with repeated assessments are needed to determine clinical utility across clinical sites and in diverse postpartum cohorts. Cutoff scores for CB-PTSD should be examined for various subgroups (e.g., degree of obstetrical complications) to optimize accuracy in screening performance. Clinician diagnostic evaluation was used as the external criterion, as the field advances, CB-PTSD biomarkers may serve as better references for test validation.

## Conclusion

Childbirth-related PTSD is an often-overlooked complication with lasting effects on mothers and infants ^12, 34^. This study shows that a brief, six-item version of the PCL-5 can reliably detect PTSD after a traumatic delivery. Because it is short, easy to score, and performs as well as the full questionnaire, it can be incorporated into routine postpartum visits and electronic health records. Widespread use of this screener would help obstetric teams recognize PTSD early, provide timely referral and support, and strengthen recovery and bonding for mothers and their babies. Further studies should confirm its performance in large and diverse populations and explore how screening can be paired with preventive or therapeutic strategies to reduce the burden of childbirth-related PTSD.

## Data Availability

All data produced in the present study are available upon reasonable request to the authors

## Declaration of generative-AI use

During the preparation of this work, the authors used ChatGPT-5 to improve language and readability and to reduce word count. The authors reviewed and edited the content as needed and take full responsibility for the final version.

## Acknowledgments

No acknowledgments to declare.

## Data availability

De-identified data available upon reasonable request and in accord with Institution’s data sharing policy.

## Authors Contribution

Alon Bartal, Ph.D.: Advised on Methodology, Formal Analysis, Software, and Visualization; Contributed to Writing – Review & Editing (Analysis and Results sections).

Hadas Allouche-Kam, M.D.: Contributed to Project Administration; Contributed to Investigation; Contributed to Visualization.

Tal Elhasid Felsenstein, M.D: Methodology, Formal Analysis and Software Elli C. Dassolpoulos, B.A.: Contributed to Investigation.

Mary C. Lee, B.S.: Contributed to Project Administration; Contributed to data curation; Contributed to Visualization.

Andrea G. Edlow, M.D.: Writing–Review and editing Scott P. Orr, Ph.D.: Writing–Review and editing

Sharon Dekel, Ph.D.: Conceptualization; Data curation; Investigation; Methodology; Resources; Supervision; Validation; Writing - original draft; Writing - Review and editing.

AB and HAK contributed equally to this work

All persons who meet authorship criteria are listed as authors, and all authors certify that they have participated sufficiently in the work to take public responsibility for the content, including participation in the concept, design, data collection, analysis, writing, or revision of the manuscript.

## Figure legends

**Figure S1 (Appendix 1).**
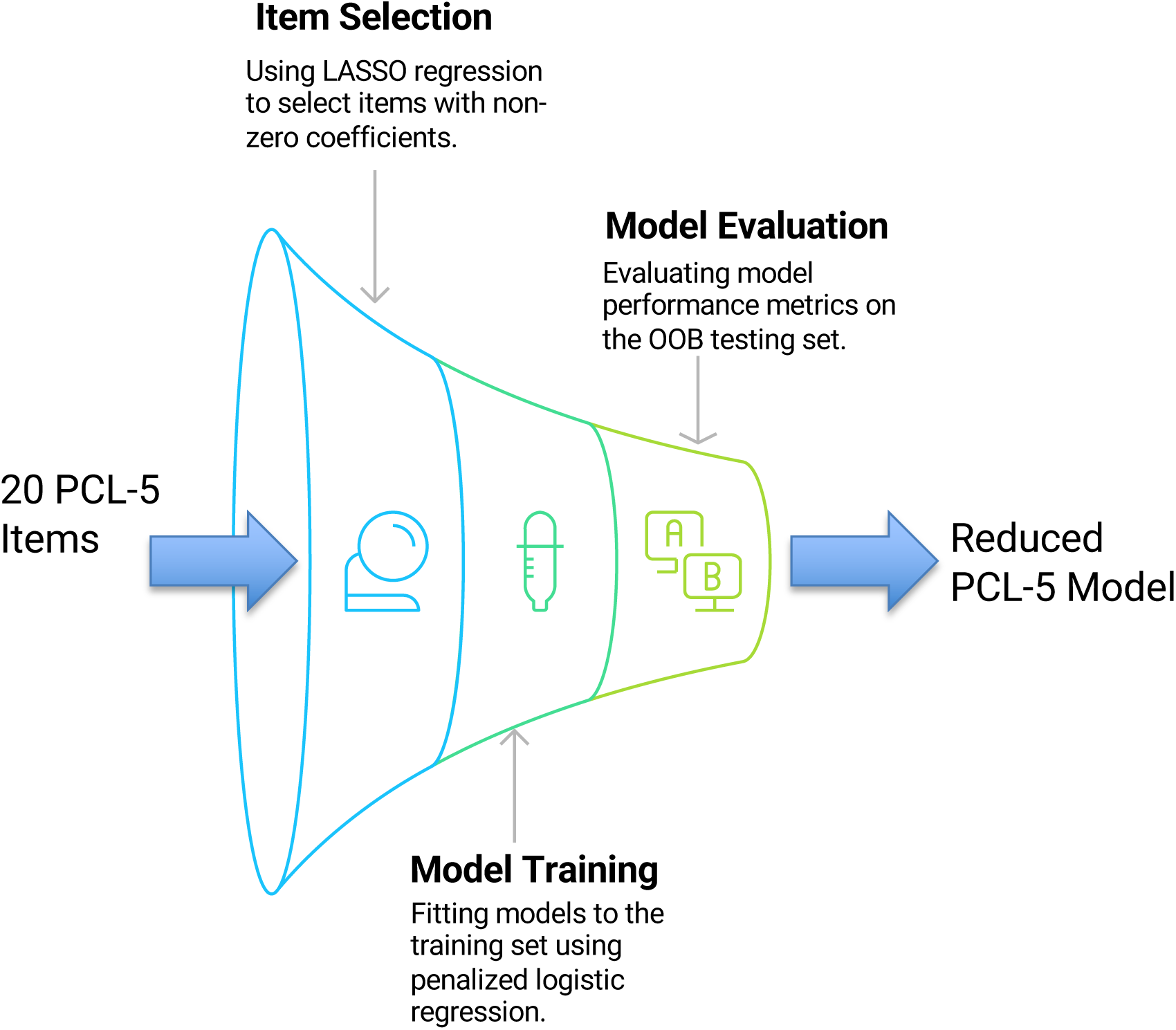
The three methodological steps for identifying the most important PTSD Checklist for DSM-5 (PCL-5) items for detecting PTSD related to childbirth.

